# Natural spring water gargle and direct RT-PCR for the diagnosis of COVID-19 (COVID-SPRING study)

**DOI:** 10.1101/2021.03.11.21251938

**Authors:** Jeannot Dumaresq, François Coutlée, Philippe J. Dufresne, Jean Longtin, Judith Fafard, Julie Bestman-Smith, Marco Bergevin, Emilie Vallières, Marc Desforges, Annie-Claude Labbé

## Abstract

We prospectively compared natural spring water gargle to combined oro-nasopharyngeal swab (ONPS) for the diagnosis of coronavirus disease 2019 (COVID-19) in paired clinical specimens (1005 ONPS and 1005 gargles) collected from 987 unique early symptomatic as well as asymptomatic individuals from the community. Using a direct RT-PCR method with the Allplex™ 2019-nCoV Assay (Seegene), the clinical sensitivity of the gargle was 95.3% (95% confidence interval [CI], 90.2 to 98.3%) and was similar to the sensitivity of the ONPS (93.8%; 95% CI, 88.2 to 97.3%), despite significantly lower viral RNA concentration in gargles, as reflected by higher cycle threshold values. No single specimen type detected all COVID-19 cases. SARS-CoV-2 RNA was stable in gargles at room temperature for at least 7 days. The simplicity of this sampling method coupled with the accessibility of spring water are clear advantages in a pandemic situation where testing frequency, turnaround time and shortage of consumables and trained staff are critical elements.

## Introduction

Testing massively, rapidly and frequently has been considered one of the most important measure to control the COVID-19 pandemic (*1, 2*). Although nasopharyngeal swab has long been considered the specimen of choice for the diagnosis of respiratory viral infections, including SARS-CoV-2 infection, it suffers from several drawbacks: its discomfort limits screening acceptability, and it is vulnerable to shortages in both specialized materials and trained healthcare workers in the context of a pandemic (*3*).

Saliva has been well studied and is now considered an acceptable alternative specimen for the diagnosis of COVID-19 (*4, 5*). However, since direct SARS-CoV-2 RT-PCR (i.e. without prior RNA extraction) has been used to accelerate turn-around time and to circumvent the problem of extraction reagent shortages, saliva may be less suitable because it usually requires RNA extraction or additional sample preparation, such as proteinase K treatment and dilution, before PCR analysis (*6*). Moreover, its viscosity can hamper laboratory automation (*7*).

Swish and gargle with a mouth rinse has been less studied as a sample for COVID-19 diagnosis. However, it is easier to obtain from individuals having difficulty producing saliva, and is also easier to manipulate in the laboratory since the sample is already diluted, which effectively reduces sample viscosity. One small study found gargle to be as sensitive as throat swab for the detection of respiratory pathogens but did not compare it to nasopharyngeal swab (*8*). Some authors have used saline as a mouth rinse and gargle and found it to be at least as sensitive as nasopharyngeal swab for the diagnosis of COVID-19 (*9-11*), but they proceeded to RNA extraction as saline is less suitable for direct RT-PCR due to inhibition of some PCR assays by high concentration of sodium chloride (*12*). Moreover, many medical grade supplies are prone to shortages during a pandemic. These shortcomings of actual studies justified our evaluation of the swish and gargle with natural spring water compared to combined oro-nasopharyngeal swab (ONPS) for the diagnosis of COVID-19 using a direct RT-PCR method without RNA extraction.

## Methods

### Study subjects and specimen collection

Patients aged ≥ 6 years presenting to a drive-through test center were included either if they had symptoms suggestive of COVID-19 or if they were considered an asymptomatic close contact by public health services. Patients aged 5 years or less were excluded because there are usually unable to gargle adequately. After obtaining consent, an ONPS was first collected using a standardised method by a trained health care professional. The flocked swab was first used to swab the posterior oropharynx and the tonsillar arches, and then the same swab was inserted through one nostril parallel to palate until resistance was met or the distance was equivalent to the distance from the patient’s ear to their nostril, rotated several times and left in place for 5 – 10 seconds prior to being removed as per Center for Disease Control instructions for collection (*13*). The ONPS was placed in a conical 15 ml centrifuge tube containing 3 ml of molecular water (PCR grade water), which is a validated, standard-of-care specimen transport medium for SARS-CoV-2 testing (*12, 14*). Subjects were then provided with 5 ml of natural spring water (Eska water, St-Mathieu-d’Harricana, Québec, Canada or Naya water, Mirabel, Québec, Canada) in a disposable soft plastic cup (Plastic Medicine Cups, AMG Medical, Montreal, Quebec, Canada) and instructed to rinse their mouth for 5 seconds, tilt their head back and gargle for 5 seconds, repeat this cycle once, expel the water back in the plastic cup and empty it in a 15 ml conical polypropylene centrifuge tube. Both specimens were simultaneously submitted and processed for SARS-CoV-2 testing by a direct one-step reverse transcription-PCR (RT-PCR) method as described below. The specimens were refrigerated at the collection site and transported to the laboratory in insulated coolers with ice packs. Detailed instructions for the gargle procedure are included in the supplemental material. This research was considered by our research ethics committee at *CISSS de Chaudière-Appalaches* and deemed exempt in accordance with the *Tri-Council Policy Statement: Ethical Conduct for Research Involving Humans – TCPS 2 (2018)*. The research was conducted in accordance with the Declaration of Helsinki.

### SARS-CoV-2 detection

All samples were vortexed for 2-3 seconds, heat inactivated at 70 °C for 10 minutes in a water bath, and centrifuged at 3000 x g for 2 minutes. ONPS transport medium (molecular grade water) was diluted 1:3 in molecular grade water (500 µl in 1500 µl) in a 5 ml cryovial tube (Simport Scientific, Beloeil, Québec, Canada). This dilution was routinely used to lower the proportion of invalid RT-PCR results (data not shown). Gargles were directly transferred (1 ml) undiluted in a 5 ml cryovial tube. Cryovial tubes were loaded on a Seegene STARlet IVD (Seegene, Seoul, Republic of Korea and Hamilton Company, Reno, NV, USA) for preparation of PCR microplates for direct RT-PCR without RNA extraction. Direct RT-PCR was performed using the Allplex™ 2019-nCoV Assay kit on CFX96 Touch Real-Time PCR Detection System (Bio-Rad, Hercules, CA, USA) as described by Merindol N et al. (*12*), and results interpreted using the Seegene Viewer software. Cycle thresholds (Ct) served as a relative indicator of viral load. Samples were considered to have low viral loads if less than 3 genes were detected or if the N gene Ct was equal to or greater than 35. This definition usually corresponds to a viral load of 720 copies/ml or less (data not shown). Results were considered invalid if neither the internal control nor any target gene were detected, which usually represents PCR inhibition. Discordant pairs and every pair containing at least one invalid result were also analyzed with the *RealTime* SARS-CoV-2 assay (Abbott, Chicago, IL, USA).

### RNA stability study

A volunteer gargled with 5 ml of four different waters (Eska; Naya; Molecular water BP2819, Fisher Bioreagents; Sterile water for irrigation, Baxter), three times with each water. One SARS-CoV-2 PCR positive ONPS non-inactivated sample was used to spike the 12 gargle specimens (1:8 dilution). One spiked gargle from each brand of water was analyzed immediately and the two other spiked gargles from each brand of water were kept seven days at room temperature and at 4 °C, respectively, before being analyzed using the same method described above. Other waters, including tap water, were also tested to evaluate RT-PCR compatibility (see Table S1 in supplemental material).

### Statistical methods

Data were described by percentage for categorical variables and median with interquartile range (IQR) for continuous variables. Both gargle and ONPS were compared to a composite reference standard, defined as positive if either the ONPS or the gargle was positive. Online MedCalc software (https://www.medcalc.org/calc/index.php) was used to calculate sensitivity, specificity, disease prevalence, predictive values, accuracy, likelihood ratios as well as comparison of proportions (chi-square test) and their respective confidence intervals. The level of agreement was also assessed using kappa statistics. By definition, Kappa values above 0.75 indicate excellent agreement, values between 0.40 and 0.75 indicate fair to good agreement, and values below 0.40 represent poor agreement beyond chance. *P* values for the comparison of means (*t*-test for paired samples) were calculated using the Statistica software (Statsoft Inc, OK, USA). *P* values for the comparison of standard deviations (F test for variances) were calculated using the online Statistics Kingdom calculator (https://www.statskingdom.com/220VarF2.html). Box-and-whisker graphs were generated using the online Good Calculators software (https://goodcalculators.com/box-plot-maker/).

## Results

A total of 2010 paired clinical specimens (1005 ONPS and 1005 gargles) were collected from 987 unique patients between October 8^th^ and 23^rd^ 2020. The average age of study participants was 40 years (range 6 to 91 years, median 40 years), 633 (63%) were symptomatic, 244 (24%) were asymptomatic but close contacts to confirmed COVID-19 cases, and 126 (13%) were asymptomatic but working in a unit or a workplace with a COVID-19 outbreak.

As shown in **Figure 1**, 121 individuals (12%) had a PCR-positive ONPS, 123 (12.2%) had a PCR-positive gargle, and 115 (11.4%) had SARS-CoV-2 detected in both samples. There was no difference in clinical sensitivity between gargle and ONPS, with a Cohen’s kappa coefficient of 0.94 (95% CI 0.89 - 0.96). Six participants had PCR-positive ONPS and PCR-negative gargle: all 6 ONPS contained low viral loads, and 3/6 gargles were found to be weakly PCR-positive when retested with the Abbott *RealTime* SARS-CoV-2 assay, which includes an RNA extraction step. Eight patients had PCR-positive gargle and PCR-negative ONPS: 6/8 gargles contained low viral loads, and 2/7 ONPS were found to be PCR-positive when retested with the Abbott *RealTime* SARS-CoV-2 assay (one ONPS was lost so could not be retested). Both types of samples showed a better clinical sensitivity with symptomatic patients, but the difference was only statistically significant when comparing gargle to the composite reference standard with discrepant analysis (**Figure 1**).

**Figure 1.**
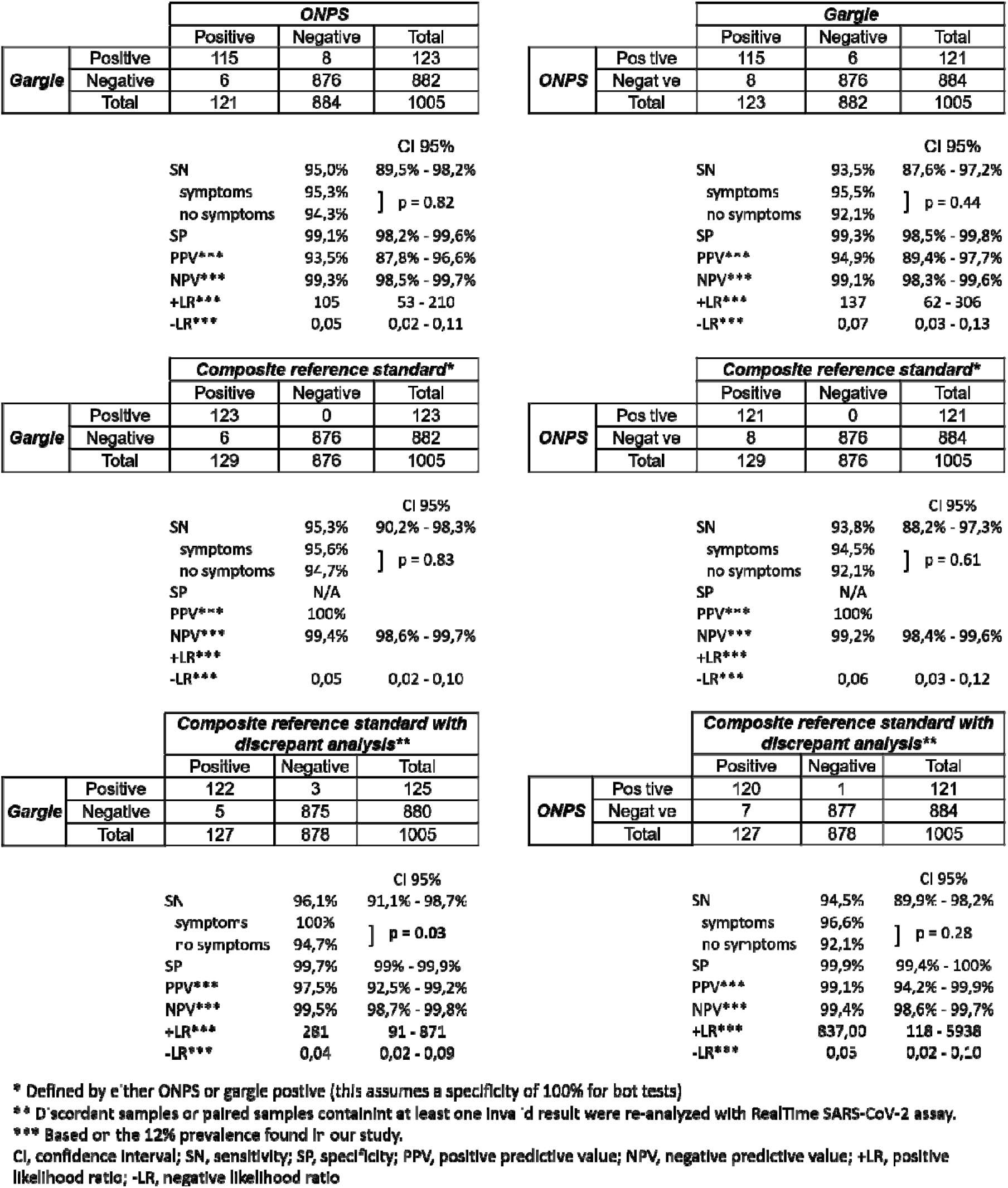
2×2 tables comparing gargle and ONPS, with composite reference standard and discrepant analysis

Results were invalid for 2.7% (27 of 1005) ONPS and for 0.9% (9 of 1005) gargles (difference 1.8%, 95% CI 0.64% to 3.07%, p=0.002).

Cycle threshold (Ct) differences were observed for concordant positive paired samples (**Figure 2**). Overall, gargles had higher Ct values with mean differences of 5.3, 5.6 and 5.2 for E, RdRp and N genes respectively (p < 0.0001). When analyzing Ct differences by different ONPS gene E Ct intervals, the differences were only significant when the ONPS had a Ct value lower than 25, representing higher viral loads (**Figure 2**). Also, there was a statistically significant difference between standard deviations of gargles and ONPS Ct values for RdRp gene (4.18 vs 5.29, p = 0.02) and N gene (4.8 vs 6.0, p = 0.01), but not E gene (4.77 vs 5.57, p = 0.09).

**Figure 2.**
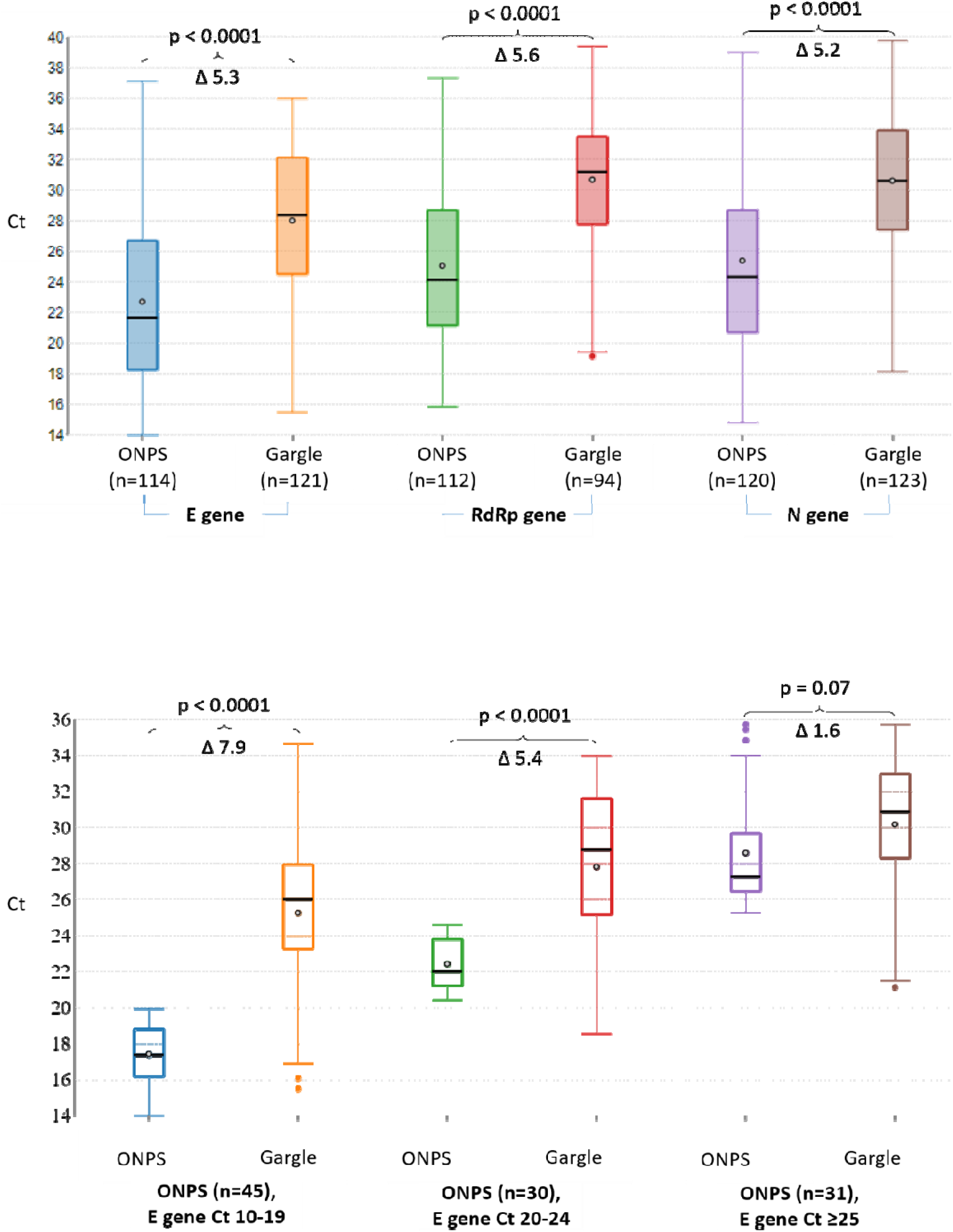
Mean Ct difference between ONPS and gargle, overall and separated by different ONPS E gene Ct intervals for concordant positive sample pairs

SARS-CoV-2 RNA was stable in natural spring waters and in gargle for at least 7 days, either at room temperature or refrigerated at 4 °C (**Table 1**).

**Table 1.** SARS-CoV-2 RNA stability in different waters and gargles.

|  | Day 0 |  |  |  | Day 7 - 4 degrees Celsius |  |  |  | Day 7 - Room temperature |  |  |  |
| --- | --- | --- | --- | --- | --- | --- | --- | --- | --- | --- | --- | --- |
|  | E gene (Ct) | RdRp gene (Ct) | N gene (Ct) | IC (Ct) | E gene (Ct) | RdRp gene (Ct) | N gene (Ct) | IC (Ct) | E gene (Ct) | RdRp gene (Ct) | N gene (Ct) | IC (Ct) |
| Naya water gargle | N/A | N/A | N/A | 26,62 |  |  |  |  |  |  |  |  |
| Naya water gargle spiked | 26,38 | 29,26 | 28,62 | 26,27 | 26,90 | 29,47 | 28,44 | 26,41 | 27,76 | 29,37 | 29,67 | 27,04 |
| **Delta** |  |  |  |  | **0,52** | **0,21** | **-0,18** | 0,14 | **1,38** | **0,11** | **1,05** | 0,77 |
| Molecular water gargle | N/A | N/A | N/A | 26,80 |  |  |  |  |  |  |  |  |
| Molecular water gargle spiked | 26,40 | 27,80 | 27,29 | 25,88 | 26,56 | 29,39 | 28,67 | 26,06 | 26,56 | 29,29 | 28,74 | 26,21 |
| **Delta** |  |  |  |  | **0,16** | **1,59** | **1,38** | 0,18 | **0,16** | **1,49** | **1,45** | 0,33 |
| Eska water gargle | N/A | N/A | N/A | 26,26 |  |  |  |  |  |  |  |  |
| Eska water gargle spiked | 26,93 | 30,19 | 29,11 | 26,06 | 27,33 | 30,74 | 29,75 | 26,56 | 27,28 | 29,70 | 29,74 | 27,23 |
| **Delta** |  |  |  |  | **0,40** | **0,55** | **0,64** | 0,50 | **0,35** | **-0,49** | **0,63** | 1,17 |
| Baxter medical water gargle | N/A | N/A | N/A | 27,81 |  |  |  |  |  |  |  |  |
| Baxter medical water gargle spiked | 27,28 | 31,26 | 29,57 | 27,29 | 27,20 | 32,35 | 29,53 | 26,97 | 27,40 | 30,97 | 30,09 | 27,23 |
| **Delta** |  |  |  |  | **-0,08** | **1,09** | **-0,04** | -0,32 | **0,12** | **-0,29** | **0,52** | -0,06 |

## Discussion

Testing frequency is considered important to control the COVID-19 pandemic, but this requires sampling methods with high acceptability in order to address testing hesitancy. Non-invasive samples like saliva or gargle clearly increase the acceptability of testing (*10*) and alleviate pressure on limited resources that are currently used to collect oro-nasopharyngeal swabs. We have gathered after this study many qualitative data confirming the impact of gargle on testing acceptability and we saw an important increase in healthcare workers adherence to systematic screening (data not shown). Gargle also widens the possibility of self-collection as it was shown to have similar sensitivity to a nasopharyngeal swab (*10, 11*). Self-collected non-invasive specimens can eliminate socio-economical barriers to testing hesitancy and increase patient satisfaction. Notwithstanding that some evidence suggests that testing frequency could be more important than analytical sensitivity (*1, 15*), it is essential to demonstrate that novel sampling methods retain acceptable sensitivity. Like others, we were able to demonstrate that gargle can be as sensitive as ONPS (*9-11*), even without RNA extraction. Our data also demonstrates that it is important to use a composite reference standard when sampling methods offer similar analytical and clinical performance to avoid underestimation of the novel sampling method (*5*). There was no difference in sensitivity between symptomatic and asymptomatic patients in our study, except when comparing gargle to composite reference standard combined with discrepant analysis, although we may have lacked power to demonstrate a difference.

Saliva generally requires dilution and/or enzymatic digestion to permit direct RT-PCR without significant rates of inhibition (*16*). Since gargle is essentially a form of pre-diluted saliva, we obtained a low rate of PCR inhibition in this study. Preliminary tests showed that some natural spring waters, as well as local tap water, did not seem to affect direct RT-PCR efficiency when compared to molecular water (see supplemental material). We chose to use commercial natural spring waters that were bottled from unique sources, which offers homogeneity and high chemical stability across time. These water sources are highly regulated and human or agricultural activity are not allowed near them. In Canada, natural spring waters are readily available and represent a cheap alternative to more expensive medical grade products that are prone to shortages during a pandemic. Moreover, we were able to demonstrate stability of viral RNA for at least 7 days at room temperature, similarly to saliva (*17, 18*). This represents a significant advantage since getting rid of the cold-chain can help support surge capacity of SARS-CoV-2 diagnostic testing (*19*).

We observed a statistically significant difference between the viral RNA concentrations (extrapolated from PCR Ct) of both sample types, gargle having a lower relative viral load when considering all concordant pairs. Some studies have demonstrated that a lower quantity of viral RNA could be detected from saliva compared to nasopharyngeal swabs (*20-22*). The lower relative viral loads measured with Cts we observed with gargle compared to EONPs could be related to the fact that gargle is a diluted saliva sample. However, the viral RNA concentrations difference was only significant in paired samples with higher ONPS viral loads (Ct < 25). This could be in part explained by the fact that viral RNA declines more rapidly in the nasopharynx, since some studies showed that SARS-CoV-2 RNA could be amplified from saliva for longer periods (*20-22*) and another study found that gargle was more often positive than ONPS even when symptoms were present for more than 7 days (*10*). We could also hypothesize that the viral RNA concentration in the throat and saliva, and thus gargle, is possibly less variable from one day or from one sample to another compared to the nasopharyngeal swab. As a matter of fact, when comparing positive gargles and ONPS our data shows statistically significant differences between the standard deviations of RdRp and N genes Cts, gargles’ Cts being less variable than those of ONPS. Moreover, some authors found greater variation in human RNase P Ct values in nasopharyngeal swab specimens than in saliva specimens (*21*), consistent with more variable nasopharyngeal swab specimens quality. The pharynx being at the junction between the upper and the lower respiratory tract, it could reflect the viral RNA content of both anatomical compartments as the virus progressively migrate from one site to another. The absence of significant Ct difference in paired samples containing lower viral loads in ONPS (Ct ≥ 25) is not explained by discordant samples, since the latter were well balanced between both sample types (6 gargle-/ONPS+ and 8 gargle+/ONPS-). One study comparing the performance of oropharyngeal swab with nasopharyngeal swab for the diagnosis of COVID-19 also found a similar sensitivity between both sample types, with discordant pairs being also well balanced between both sample types, and yet they found higher Ct values in oropharyngeal swabs (*23*).

Our study has some limitations. Since we were using a direct RT-PCR method, ONPS transport medium had to be diluted 1:3 to avoid PCR inhibition and hence decrease invalid results rate.

This dilution could have decreased ONPS clinical sensitivity at the advantage of gargle. Also, symptomatic patients in this study were all recruited within the first seven days of symptom onset so we cannot extrapolate our results to patients presenting later in the disease course. Moreover, we cannot either extrapolate our results to nursing home patients or infants/toddlers who were not included in this study, and who, in our experience, provide gargle samples with more difficulty.

## Conclusion

Natural spring water gargle is as sensitive as ONPS for the diagnosis of SARS-CoV-2 in early symptomatic as well as asymptomatic patients from the community using a direct RT-PCR method with the Allplex™ 2019-nCoV assay. Non-invasive sampling methods will increase acceptability of SARS-CoV-2 testing and reduce the need for precious medical grade products and trained healthcare workforce. More studies are needed and are underway to evaluate the clinical performance of natural spring water gargle with other PCR assays for the diagnosis of COVID-19.

## Data Availability

All data analysed during this study are included in this published article and are available from the authors upon reasonable request.

## Acknowledgements

The authors report no potential conflicts of interest relevant to this manuscript. This study was supported by the Ministère de la Santé et des Service sociaux de la province de Québec (reagents and equipment) and the Centre intégré de santé et de services sociaux de Chaudière-Appalaches (human and technical resources). We would particularly like to thank nurses and laboratory technologists from the hospital Hôtel-Dieu de Lévis, all the staff from the community assessment centers Archimède and Paul-Gilbert, as well as all the participants.

## Supplemental material

**Table S1.** Different waters evaluated for RT-PCR compatibility.

|  | E gene (Ct) | RdRp gene (Ct) | N gene (Ct) | IC (Ct) |
| --- | --- | --- | --- | --- |
| Naya water (pure) | N/A | N/A | N/A | 26,12 |
|  | N/A | N/A | N/A | 25,91 |
| Eska water (pure) | N/A | N/A | N/A | 26,24 |
|  | N/A | N/A | N/A | 25,51 |
| Labrador water (pure) | N/A | N/A | N/A | 26,16 |
|  | N/A | N/A | N/A | 25,42 |
| Nestlé water (pure) | N/A | N/A | N/A | 27,94 |
| Baxter medical water (pure) | N/A | N/A | N/A | 26,12 |
|  | N/A | N/A | N/A | 25,49 |
| AddiPak sterile water (pure) | N/A | N/A | N/A | 25,47 |
| Hospital tap water (pure) | N/A | N/A | N/A | 25,39 |
| Naya water (spiked) | 29,91 | 31,91 | 32,22 | 25,36 |
| Eska water (spiked) | 29,69 | 31,47 | 31,73 | 25,38 |
| Labrador water (spiked) | 29,77 | 31,81 | 32,45 | 25,42 |
| Baxter medical water (spiked) | 29,94 | 31,55 | 32,08 | 25,27 |
| AddiPak sterile water (spiked) | 29,19 | 31,20 | 31,59 | 25,35 |
| Hospital tap water (spiked) | 29,72 | 32,18 | 31,88 | 25,35 |

## Notes

### Competing Interest Statement

The authors have declared no competing interest.

### Funding Statement

This study was supported by the Ministere de la Sante et des Service sociaux de la province de Quebec (reagents and equipment) and the Centre integre de sante et de services sociaux de Chaudiere-Appalaches (human and technical resources).

### Author Declarations

This research was considered by our research ethics committee at CISSS de Chaudiere-Appalaches and deemed exempt in accordance with the Tri-Council Policy Statement: Ethical Conduct for Research Involving Humans - TCPS 2 (2018). The research was conducted in accordance with the Declaration of Helsinki.

